# Bimodal distribution and set point HBV DNA viral loads in chronic infection: retrospective analysis of cohorts from the UK and South Africa

**DOI:** 10.1101/2020.01.08.20016972

**Authors:** Louise O Downs, Sabeehah Vawda, Phillip Armand Bester, Katrina A Lythgoe, Tingyan Wang, Anna L McNaughton, David A Smith, Tongai Maponga, Oliver Freeman, Kinga A Várnai, Jim Davies, Kerrie Woods, Christophe Fraser, Eleanor Barnes, Dominique Goedhals, Philippa C Matthews

**Affiliations:** Department of Infectious Diseases and Microbiology, Oxford University Hospitals NHS Foundation Trust, Oxford, UK; Nuffield Department of Medicine, Medawar Building for Pathogen Research, University of Oxford, Oxford, UK; Division of Virology, University of the Free State and National Health Laboratory Service, Bloemfontein, South Africa; Big Data Institute, Li Ka Shing Centre for Health Information and Discovery, Nuffield Department of Medicine, University of Oxford, United Kingdom; Department of Zoology, University of Oxford, M Medawar Building for Pathogen Research, Nuffield Department of Medicine, University of Oxford, Oxford, UK; National Institute of Health Research Health Informatics Collaborative, NIHR Oxford Biomedical Research Centre, Oxford, UK; Department of Virology, University of Stellenbosch, Tygerberg Hospital, Cape Town, South Africa; Department of Hepatology, Oxford University Hospitals NHS Foundation Trust, Oxford, UK; Oxford University Hospitals NHS Foundation Trust, Oxford, UK; Department of Computer Science, University of Oxford, Oxford, UK; Nuffield Department of Population Health, University of Oxford, Oxford, UK

## Abstract

Hepatitis B virus (HBV) DNA viral loads (VL) show wide variation between individuals with chronic hepatitis B (CHB) infection, and are used to determine treatment eligibility. There are few refined descriptions of VL distribution in CHB, and limited understanding of the biology that underpins these patterns. We set out to describe the VL distribution in independent cohorts from the UK and South Africa, to identify associated host characteristics, and to compare with VL in HIV-1 and hepatitis C (HCV) infection. We show the presence of a set-point in chronic HBV infection and report significant differences in the viral load distribution of HBV when compared to HIV and HCV.

## Introduction

Hepatitis B virus (HBV) DNA viral loads (VL) show wide variation between individuals with chronic hepatitis B (CHB) infection, and are used to determine treatment eligibility [1–3]. The relationship between HBV e-antigen (HBeAg)-positive status and high VL in CHB is well recognised [4], but there are few refined descriptions of VL distribution, and limited understanding of the biology that underpins these patterns. Set point viral load (SPVL), defined as a stable level of viraemia in peripheral blood during the initial years of chronic infection, is a concept well established in HIV [5], but has not been explored for HBV infection to date.

Developing improved insights into the distribution of VL at a population level is important for planning wider treatment deployment to support progress towards international sustainable development goals for HBV elimination [6], which set ambitious targets for reducing morbidity and incidence of new CHB cases [7]. Characterisation of HBV VL dynamics is also important for mathematical modelling, and for generating new insights into persistence, transmission and pathogenesis.

We have therefore set out to describe the VL distribution in independent cohorts from the UK and South Africa, to identify associated host characteristics, to compare with VL distributions in HIV-1 and hepatitis C (HCV) infection, and to seek evidence for SPVL in HBV infection.

## Methods

We retrospectively collected VL measurements ± supporting metadata for adults with chronic HBV, HCV and HIV infection from four cohorts:

I. **HBV: UK dataset** We collected data for adults with CHB infection from electronic records at Oxford University Hospitals NHS Foundation Trust, as part of the National Institute of Health Research Health Informatics Collaborative (NIHR-HIC [8]), as previously described [9]. We assimilated VL results (Abbott M2000 platform) for 371 individuals over six years commencing 1st January 2011, for whom baseline HBeAg status was available in 351 (95%). Age, sex and self-reported ethnicity (using standard ethnicity codes) were available for 352, 355 and 322 individuals, respectively. For longitudinal VL analysis, we only used data prior to commencing antiviral treatment, including patients with ≥2 measurements (n=299 individuals, 1483 timepoints).
II. **HBV: South Africa dataset** We collected all HBV VL data from the South African National Health Laboratory Service (NHLS) recorded over a four year period commencing 1^st^ January 2015 (n=6506 individuals). These were generated using various commercial platforms in different NHLS labs across the country. Other metadata (HBeAg status, HIV status, treatment data) were not available. For the purposes of analysis, we excluded VL measurements below the limit of detection based on the assumption that the majority of these samples were taken on antiviral treatment (indicated for HBV infection ± HIV co-infection). All those above the laboratory limit of quantification were designated 1.7×10^8^ IU/ml. For analysis of longitudinal data, we included patients with ≥2 detectable VL measurements (n=874 individuals; 9578 timepoints).
III. **HCV** Baseline HCV viral loads were collected for adults prior to commencing antiviral treatment between 2006-2018, representing 925 individuals, from the same source as the UK HBV data using the Abbott M2000 platform, and collected through the NIHR-HIC pipeline [9]. The setting and characteristics of this study population has been previously described [10].
IV. **HIV** HIV data were obtained from a UK database of HIV seroconverters between 1985-2014 through the BEEHIVE collaboration (n=1581) [5]. HIV VL was measured using COBAS AmpliPrep/COBAS TaqMan HIV-1 Test, v2.0 on samples collected starting at 6-24 months after infection. SPVL was defined as the average VL for each patient over time, as previously described [5].

### Statistical analysis

We used Graphpad Prism v.8.2.1 for analysis of VL distributions, skewness, and univariate analysis of patient parameters associated with HBV VL (Mann Whitney U test and Kruskall Wallis test). HBV and HCV VL are conventionally reported in IU/ml, but to make direct comparisons between VL in different infections, we also converted data into copies/ml (1 IU = 5.4 copies/ml for HBV [11] and 2.7 copies/ml for HCV [12]).

We used R package to assess within and between patient VL variability, using longitudinal data from UK HBeAg-negative adults, and from South African individuals with detectable VL. A large contribution of between-host variation would provide support for SPVL. We defined total variation, between-individual and within-individual variation according to analysis of variance (ANOVA). Specifically, the calculations are as follows:

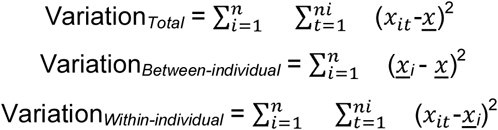

*n* denotes the number of individuals; *n*_*i*_ represents the number of data points for individual *i*; *x*_*it*_ denotes the viral load of patient *i* at time point *t*; *<u>x</u>* is the mean of viral loads of all data points; *<u>x</u>*_*it*_ is the mean of viral loads of patient *i*.

### Ethics

Data collection for the UK cohort was approved by the NRES Committee South Central-Oxford C (ref: 15/SC/0523). South African data collection was approved by the Health Sciences Research Ethics Committee at the University of the Free State (ref: UFS-HSD2019/0044/2603).

### Data Availability

Suppl Fig 1 and full metadata for our study cohorts are available on-line at figshare, DOI: 10.6084/m9.figshare.11365082.

## Results

Our UK HBV cohort was 56% male, median age 42 years, with diverse ethnic backgrounds (among 322 individuals with self-reported ethnicity data, 38% were Asian, 34% White, 24% Black, 4% Arabic, <1% other). Overall, median HBV VL was 3.4 log_10_ IU/mL; 95% CI 3.2 – 3.5 log_10_ IU/mL (equivalent to median 4.2 log_10_ copies/ml). There was a bimodal VL distribution with two peaks:

**Figure 1:**
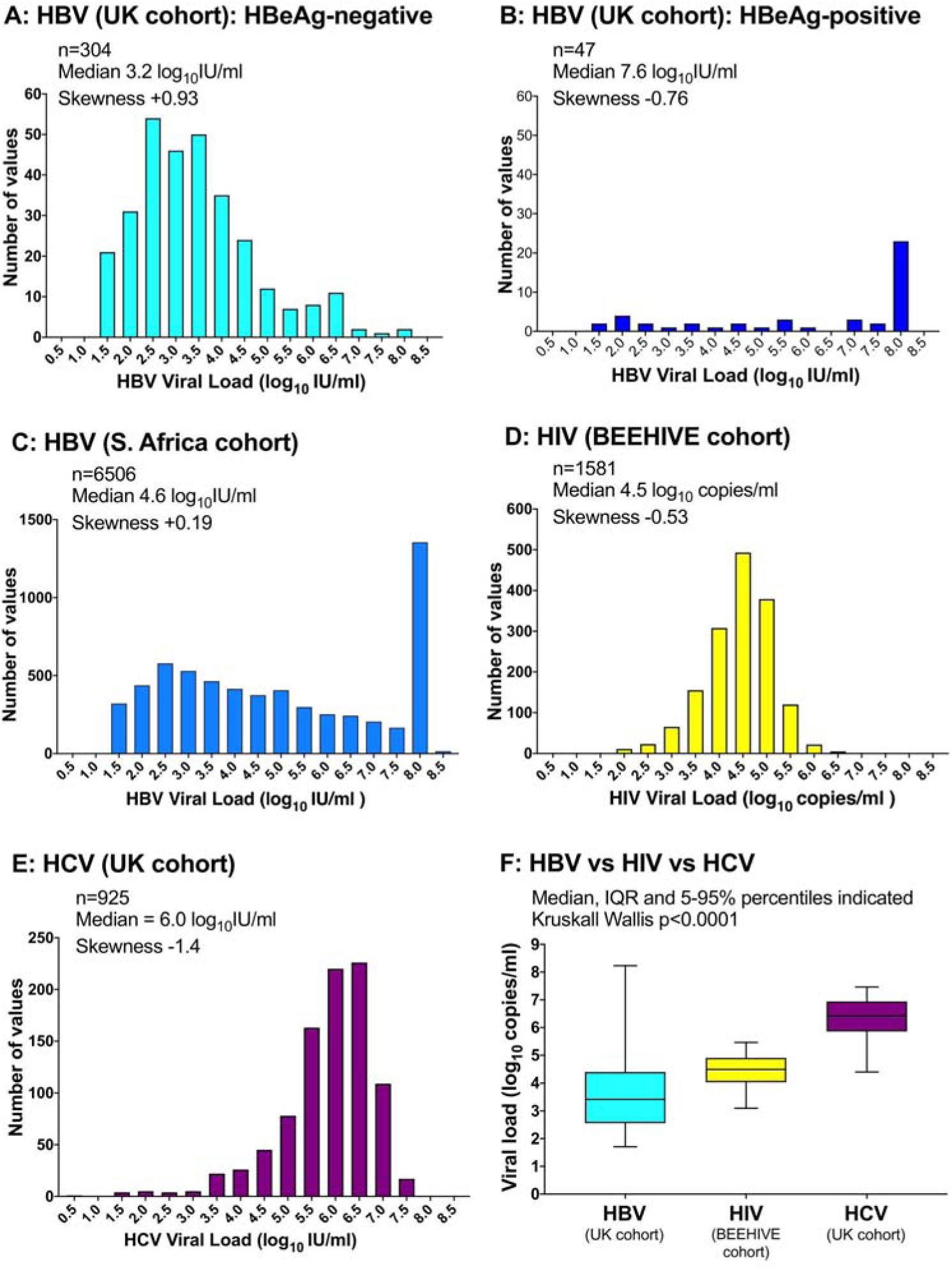
Distribution of viral loads for adults with chronic infection with HBV, HIV and HCV.

i. **HBeAg-negative infection** (accounting for 304/351 (87%) of measurements): median VL 3.2 log_10_ IU/mL (95% CI 3.0 – 3.4); right-skewed distribution (Fig 1A);
ii. **HBeAg-positive infection** (accounting for 47/351 (13%) of measurements): median VL 7.6 log_10_ IU/mL (95% CI 5.6 – 8.2); left-skewed distribution (Fig 1B).

In the South African dataset (HBeAg status not determined), median HBV VL was 4.6 log_10_ IU/mL (95% CI 3.9 – 4.0), with a bimodal distribution and right-skew (Fig 1C). Median HIV VL was 4.5 log_10_ copies/mL and median HCV VL was 6.0 log_10_ IU/ml (6.4 log_10_ copies/ml), with a left skew and no bimodal distribution (Fig 1D,E). Median viraemia was significantly different between HBV, HIV and HCV (p<0.0001; Fig 1F).

For the UK data we investigated whether sex, age or ethnicity had any influence on VL; the only significant association was lower VL with increasing age in the HBeAg-positive group (p=0.01 by Kruskal Wallis, Suppl. Fig 1).

Inter-patient variation accounted for 82.7% and 88.0% of the variability in UK and South African longitudinal datasets respectively, whilst within-patient variation accounted for 17.3% and 12.0%. This provides support for a stable SPVL within individuals with CHB.

## Discussion

We report a consistent bimodal distribution of VL in CHB in a diverse UK population and a large South African dataset, in keeping with previously published studies (e.g. [13–15]), and reflecting the role of HBeAg in immunomodulation [16]. However, descriptions of this pattern have not previously been carefully refined.

HBV viral loads in HBeAg-negative infection are significantly lower than HCV and HIV, which may relate to differences in viral population structure, viral fitness, host immune responses, and the availability of target cells. These factors might also explain why HIV, HCV and HBeAg-positive infection have left skew VL distributions, whereas HBeAg-negative infection has a right skew. This is the first study to demonstrate the concept of SPVL in HBV infection,with between-host factors explaining >80% of the variation in VL during HBeAg-negative CHB. Missing metadata is a limitation for further analysis of our South African dataset, and longer term aspirations will be to investigate larger VL datasets together with more robust longitudinal clinical and laboratory data.

Whole genome sequencing has the potential to increase our understanding of HBV but approximately 50% of cases fall below the current sequencing threshold [17]. This means that at present there is a significant ‘blind spot’ in sequence data, preventing analysis of sequence variants in individuals with VL below the population median. Enhanced descriptions of HBV VL may shed light on the biology of chronic HBV infection, inform mathematical models of viral population dynamics within and between hosts, improve understanding of risk factors for transmission and disease progression, underpin optimisation of viral sequencing methods, and help to stratify patients for clinical trials and treatment.

## Data Availability

Suppl Fig 1 and full metadata for our study cohorts are available on-line at figshare.

https://doi.org/10.6084/m9.figshare.11365082

---

**Suppl Fig 1: Relationship between hepatitis B viral load, HBeAg status and (A) sex, (B) age, and (C) ethnicity, in a cohort of adults with chronic hepatitis B virus infection recruited in Oxford, UK**. Available on-line, DOI: 10.6084/m9.figshare.11365082

## AUTHORS’ CONTRIBUTIONS

- Study concept and design: PCM, ALM.
- Ethical approvals: PCM, EB, SV, DG
- Development of the health informatics (HIC) pipeline: DAS, JD, OF, KV, KW, EB
- Acquisition and interpretation of South African data: SV, PAB, TM, DG, LOD
- Acquisition and interpretation of UK data: LOD, DAS, OF, ALM
- Analysis of the data: LOD, TW, CF, KAL
- Wrote the manuscript: LOD, PCM
- Critical revision of manuscript: ALM, KAL
- Edited and approved the final manuscript: All authors

